# Bayesian Interim Analysis and Efficiency of Phase III Randomized Trials

**DOI:** 10.1101/2024.06.27.24309608

**Authors:** Alexander D. Sherry, Pavlos Msaouel, Avital M. Miller, Timothy A. Lin, Gabrielle S. Kupferman, Joseph Abi Jaoude, Ramez Kouzy, Molly B. El-Alam, Roshal Patel, Alex Koong, Christine Lin, Tomer Meirson, Zachary R. McCaw, Ethan B. Ludmir

## Abstract

**IMPORTANCE:** Improving the efficiency of interim assessments in phase III trials should reduce trial costs, hasten the approval of efficacious therapies, and mitigate patient exposure to disadvantageous randomizations.

**OBJECTIVE:** We hypothesized that *in silico* Bayesian early stopping rules improve the efficiency of phase III trials compared with the original frequentist analysis without compromising overall interpretation.

**DESIGN:** Cross-sectional analysis.

**SETTING:** 230 randomized phase III oncology trials enrolling 184,752 participants.

**PARTICIPANTS:** Individual patient-level data were manually reconstructed from primary endpoint Kaplan-Meier curves.

**INTERVENTIONS:** Trial accruals were simulated 100 times per trial and leveraged published patient outcomes such that only the accrual dynamics, and not the patient outcomes, were randomly varied.

**MAIN OUTCOMES AND MEASURES:** Early stopping was triggered per simulation if interim analysis demonstrated ≥ 85% probability of minimum clinically important difference/3 for efficacy or futility. Trial-level early closure was defined by stopping frequencies ≥ 0.75.

**RESULTS:** A total of 12,451 simulations (54%) met early stopping criteria. Trial-level early stopping frequency was highly predictive of the published outcome (OR, 7.24; posterior probability of association, >99.99%; AUC, 0.91; *P* < 0.0001). Trial-level early closure was recommended for 82 trials (36%), including 62 trials (76%) which had performed frequentist interim analysis. Bayesian early stopping rules were 96% sensitive (95% CI, 91% to 98%) for detecting trials with a primary endpoint difference, and there was a high level of agreement in overall trial interpretation (Bayesian Cohen’s κ, 0.95; 95% CrI, 0.92 to 0.99). However, Bayesian interim analysis was associated with >99.99% posterior probability of reducing patient enrollment requirements (*P* < 0.0001), with an estimated cumulative enrollment reduction of 20,543 patients (11%; 89 patients averaged equally over all studied trials) and an estimated cumulative cost savings of 851 million USD (3.7 million USD averaged equally over all studied trials).

**CONCLUSIONS AND RELEVANCE:** Bayesian interim analyses may improve randomized trial efficiency by reducing enrollment requirements without compromising trial interpretation. Increased utilization of Bayesian interim analysis has the potential to reduce costs of late-phase trials, reduce patient exposures to ineffective therapies, and accelerate approvals of effective therapies.

**KEY POINTS:** *Question:* What are the effects of Bayesian early stopping rules on the efficiency of phase III randomized oncology trials?

*Findings:* Individual-patient level outcomes were reconstructed for 184,752 patients from 230 trials. Compared with the original interim analysis strategy, *in silico* Bayesian interim analysis reduced patient enrollment requirements and preserved the original trial interpretation.

*Meaning:* Bayesian interim analysis may improve the efficiency of conducting randomized trials, leading to reduced costs, reduced exposure of patients to disadvantageous treatments, and accelerated approval of efficacious therapies.

## INTRODUCTION

Phase III randomized trials are designed to provide practice-changing answers to contemporary research questions. With this intent, these trials require considerable investments of many patients and their families who volunteer to participate. Years to decades of accrual and follow-up, often spanning dozens of institutions and multiple countries, can drive the costs of any one trial well beyond millions of USD.^1, 2^

Identification of more efficient mechanisms for clinical trial conduct remains an imperative for reducing the costs of completing a trial, allowing more trials to be conducted, sparing patients from exposure to disadvantageous therapies, and bringing effective new therapies to patients sooner.^3–5^ Accordingly, it is a standard practice to conduct interim analyses to determine whether a phase III study can be closed early for efficacy or futility.^4^ However, these interim analyses are usually based on *P* value calculations, which are influenced by the number of analyses performed and can fluctuate between statistical significance and non-significance with small perturbations to the underlying data.^4, 6–9^ In contrast, Bayesian analyses incorporate prior information to directly inform probability calculations at subsequent assessments, which may improve the robustness of interim assessments, and have more intuitive interpretations than *P* values.^10–16^ These and other properties of Bayesian statistics, including the ability to provide both refutational evidence (against the null hypothesis) and confirmational evidence (for the null hypothesis), are particularly appealing for interim analyses.^7, 17^

We therefore hypothesized that the addition of Bayesian early stopping rules to randomized phase III trials would improve efficiency without compromising the overall trial interpretation compared to the original analysis.^18, 19^ To test this hypothesis, we manually reconstructed 184,752 individual patient outcomes from 230 phase III oncology trials and leveraged those outcomes to study the effects of *in silico* Bayesian interim analysis following accrual simulations under various conditions.

## METHODS

### Inclusion and exclusion criteria

Phase III oncology trials were screened on ClinicalTrials.gov for this cross-sectional study. Superiority, two-arm trials with a published time-to-event primary endpoint (PEP) including mature Kaplan-Meier curves with the number of patients at risk were assessed for reconstruction. WebPlotDigitizer was used to reconstruct individual patient-level data from each PEP Kaplan-Meier curve of eligible trials.^20, 21^ Reconstruction quality was assessed by first computing a hazard ratio (HR) on the reconstructed data and then determining the absolute value of the natural logarithm of the ratio comparing the reconstructed and published HRs (i.e., abs(ln(HR_reconstructed_/HR_reported_))). High quality was defined by an absolute log ratio ≤ 0.1. Trials with low reconstruction quality were excluded. Trials were also excluded if the proportional hazards assumption was not supported in the reconstructed data according to the Schoenfeld residuals test, because the Bayesian interim analysis and published analysis used a Cox proportional hazards regression. Institutional review board approval was not required due to the use of publicly available data. This study conforms with STROBE and ROBUST guidelines.^22, 23^

### Outcomes

The primary objective was to test the hypothesis of whether *in silico* Bayesian early stopping rules improve randomized trial efficiency, defined by the number of enrolled patients required, without compromising the trial interpretation (the published PEP findings after full enrollment), compared to the original analysis. We also modeled approximate cost savings of Bayesian early stopping rules by estimating per patient costs of $41,413 USD based on an independent study.^2^

### Accrual simulations

We simulated 100 partial enrollments to each trial using a grid of randomly varying accrual parameters (**Supplement**). Briefly, the timing of each patient’s enrollment was randomly assigned, governed by an accrual rate that was randomly varied between 5 to 15 patients per month. This rate was based on the estimated average accrual rates from the underlying trials, estimated as the number of enrolled patients divided by the difference in reported accrual stop and start times. This estimation yielded a median accrual rate of 15 patients per month, with an interquartile range (IQR) of 9 to 24 patients per month, also consistent with the findings of an independent study.^24^ While accrual dynamics were simulated, the survival duration and censoring status of each patient were set to the *ab origine* values observed in the published trial after the patient met the requisite follow-up time. In this way, patient fates were predetermined, not subject to simulation, and consistent with their published outcomes.

A single interim analysis was used, rather than multiple interim analyses, to strengthen the interpretation of the findings by reducing opportunities for stopping. The interim analysis was triggered by event and enrollment criteria, once either 50% of total events had been reached or two-thirds of total enrollment had been reached, whichever occurred first. An accrual hold time randomly varying between 3 to 6 months was added after either interim analysis criterion was met prior to data locking.

### Posterior probability calculation

The posterior probabilities of efficacy and futility were calculated using a Bayesian Cox proportional hazards regression with the R software package ‘brms’.^25^ The minimum clinically important difference (MCID) was defined at a HR of 0.8 on the basis of American Society of Clinical Oncology criteria.^26^ The minimum detectable effect was defined by MCID/k, where k = 3. The effect size MCID/k was chosen as the threshold of interest in lieu of the MCID itself, because a treatment effect both superior to and at the MCID would likely be well-supported as a clinically meaningful treatment. Efficacy was defined by a posterior probability ≥ 85% for a superiority minimum detectable effect [< ln(0.8)/3]. On the basis of published guidelines, a moderately skeptical prior was adopted for the symmetrized ln(HR) when calculating the posterior probability for efficacy.^27^ Specifically, the prior was a *N* (0, 0.355) distribution, where *N* (mean, standard deviation) denotes a normal distribution with the given mean and standard deviation. Futility was defined by a posterior probability ≥ 85% for the opposite effect [≥ ln(0.8)/3], which encompassed inferiority and equivalence. A moderately optimistic prior defined as *N* (−0.41, 0.40) was used for the futility posterior.^27^ Markov chain Monte Carlo sampling estimated the posterior distribution with 4 chains, 2000 steps of burn-in, and 8000 total iterations. Convergence was defined by an effective sample size ≥ 1000 and 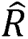 < 1.05 (here “sample size” refers to the number of independent draws from the posterior), and all models met convergence criteria.

### Statistical analysis

Continuous variables were compared with the Mann-Whitney U or Wilcoxon signed-rank tests, with statistical significance defined at *P* < 0.05. Bayesian Mann-Whitney U and Wilcoxon signed-rank tests were performed with R software package ‘dfba’ using the moderately skeptical beta prior (10, 10) to test the hypothesis that Ω or □_W_, respectively, were > 0.5 or < 0.5 (depending on the comparison). Ω and □_w_ are fundamental measures of the relative dominance between groups, where Ω or □_w_ = 0.5 indicate lack of dominance.^28, 29^ Bayes factor (BF) was also calculated, where BF > 100 represents extreme evidence of difference between groups.^30^ Multivariable Bayesian logistic regressions, fit with ‘brms’ using the same Markov chain Monte Carlo parameters as the survival models, calculated adjusted odds ratios (aOR) and 95% credible intervals (95% CrI) using the skeptical prior *N* (0, 0.355) defined on the symmetric ln(OR) scale.^25, 27^ Confounders for adjusted regressions were identified by structural causal models depicted on directed acyclic graphs using DAGitty (**Figure S1; Table S1**).^31^ Agreement in trial interpretation was defined by Bayesian Cohen’s κ, fit with the R software package ‘bfw’ using default parameters, where κ > 0.81 indicates near-perfect agreement.^32, 33^ All modeling was performed using R version 4.3.2 (Vienna, Austria).^34^ Plots were made in either R or Prism version 10 (GraphPad, La Jolla, CA).

## RESULTS

After screening, 230 trials were eligible for analysis (**Figure S2**). The PEP was OS in 90 trials (39%), and a surrogate endpoint in the remaining trials (**Table S2**). The PEP was interpreted as statistically superior in 120 trials (52%), as inferior in 5 trials (2%), and as not significantly different in 105 trials (46%). At least one frequentist interim analysis was performed in 170 trials (74%). There were no covariates associated with whether an interim analysis was performed or not. The median number of interim analyses among trials with an interim analysis was 1 (range, 1 to 11), and 64 trials (28%) were closed early on the basis of the frequentist interim analysis.

Trial accrual was halted for the *in silico* Bayesian interim analysis in 13,732 simulations (60%) after the event criterion was reached, and in the remaining 9268 simulations (40%) after the enrollment criterion was reached. A total of 12,451 simulated trials (54%) were recommended for early stopping, either on the basis of efficacy in 9988 simulated trials (80%) or futility of the experimental arm in 2463 simulated trials (20%). At the trial level, the median frequency of early stopping was 53% (IQR, 20% to 90%) (**Figure 1A**). For trials with an OS PEP or surrogate PEP, the median early stopping frequency was 33% (IQR, 17% to 65) and 71% (IQR, 26% to 100%), respectively (**Figure 1B-C**). There was >99.99% posterior probability (Ω, 0.34; 95% CrI, 0.28 to 0.41; BF_01_, 1.6 x 10^5^, by Bayesian Mann-Whitney U) for a difference in stopping frequency between trials with OS vs surrogate PEPs (*P* < 0.0001 by Mann-Whitney U) (**Figure S3**).

**Figure 1.**
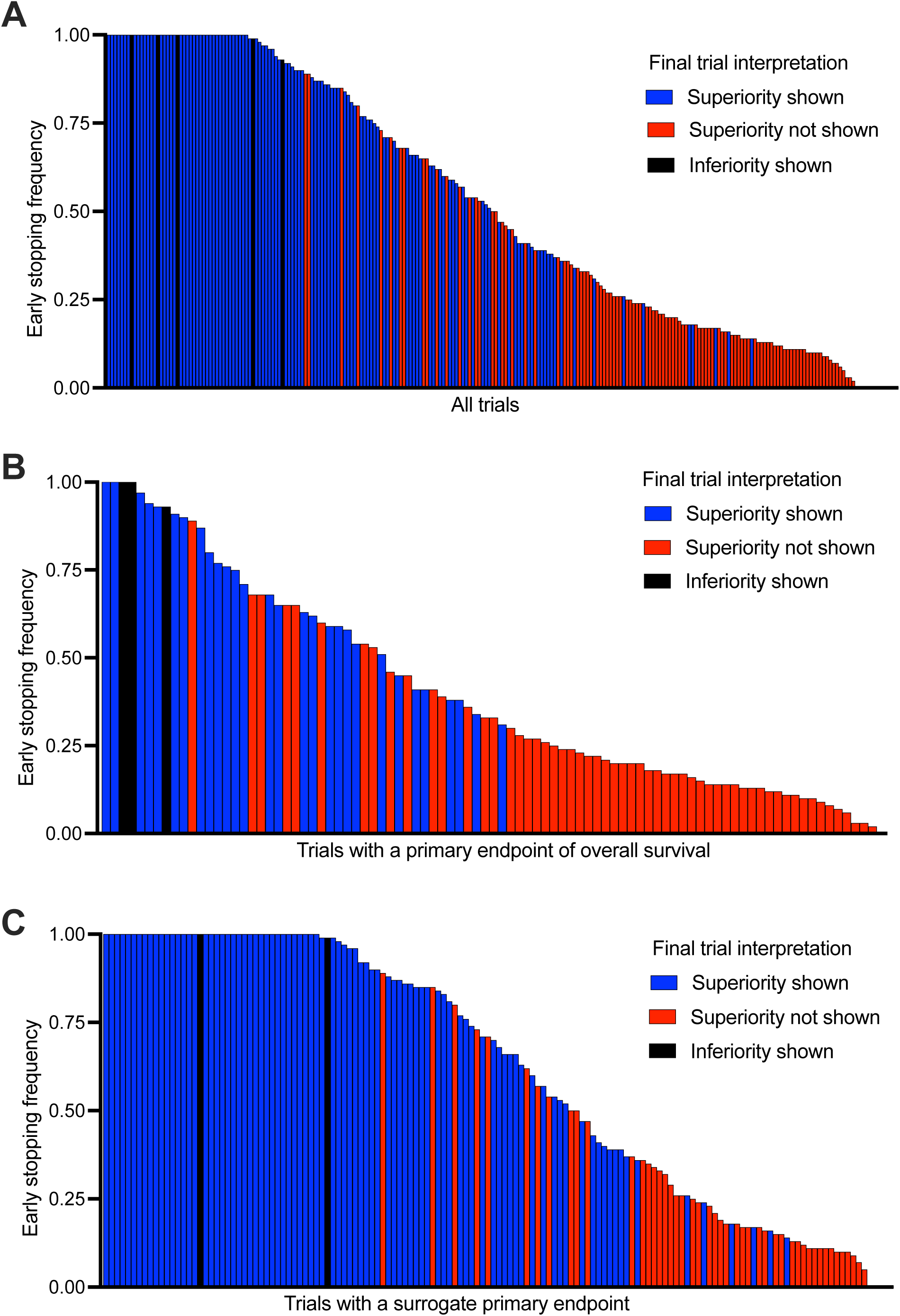
Early stopping frequency for each trial, labeled according to the final published interpretation of the primary endpoint. (A) overall dataset, (B) trials with an overall survival primary endpoint, and (C) trials with a surrogate primary endpoint.

The association between the early stopping frequency and the original trial outcome was strong (OR, 7.24; 95% CrI, 4.18 to 12.18; posterior probability of association, >99.99%). The median early stopping frequency for trials that showed differences in the PEP (i.e., either superiority or inferiority) was 87% (IQR, 59% to 100%), compared with 20% for trials without PEP differences (IQR, 13% to 35%) (*P* < 0.0001 by Mann-Whitney U; Ω, 0.88; 95% CrI, 0.83 to 0.92; BF_10_, approaching ∞, by Bayesian Mann-Whitney U) (**Figure 2A-B**). Consistent with this finding, the area under the receiver operating characteristic curve for stopping frequency and PEP difference was 0.91 (95% CI, 0.88 to 0.95, *P* < 0.0001) (**Figure S4**). The association between early stopping frequency and original outcome was similarly strong for trials with an OS PEP (*P* < 0.0001 by Mann-Whitney U; posterior probability, >99.99%; Ω, 0.82; 95% CrI, 0.74 to 0.90; BF_10_, 1.8 x 10^10^, by Bayesian Mann-Whitney U) and surrogate PEP trials (*P* < 0.0001 by Mann-Whitney U; posterior probability, >99.99%; Ω, 0.85; 95% CrI, 0.78 to 0.91; BF_10_, 9.0 x 10^15^, by Bayesian Mann-Whitney U) (**Figure 2C-F**).

**Figure 2.**
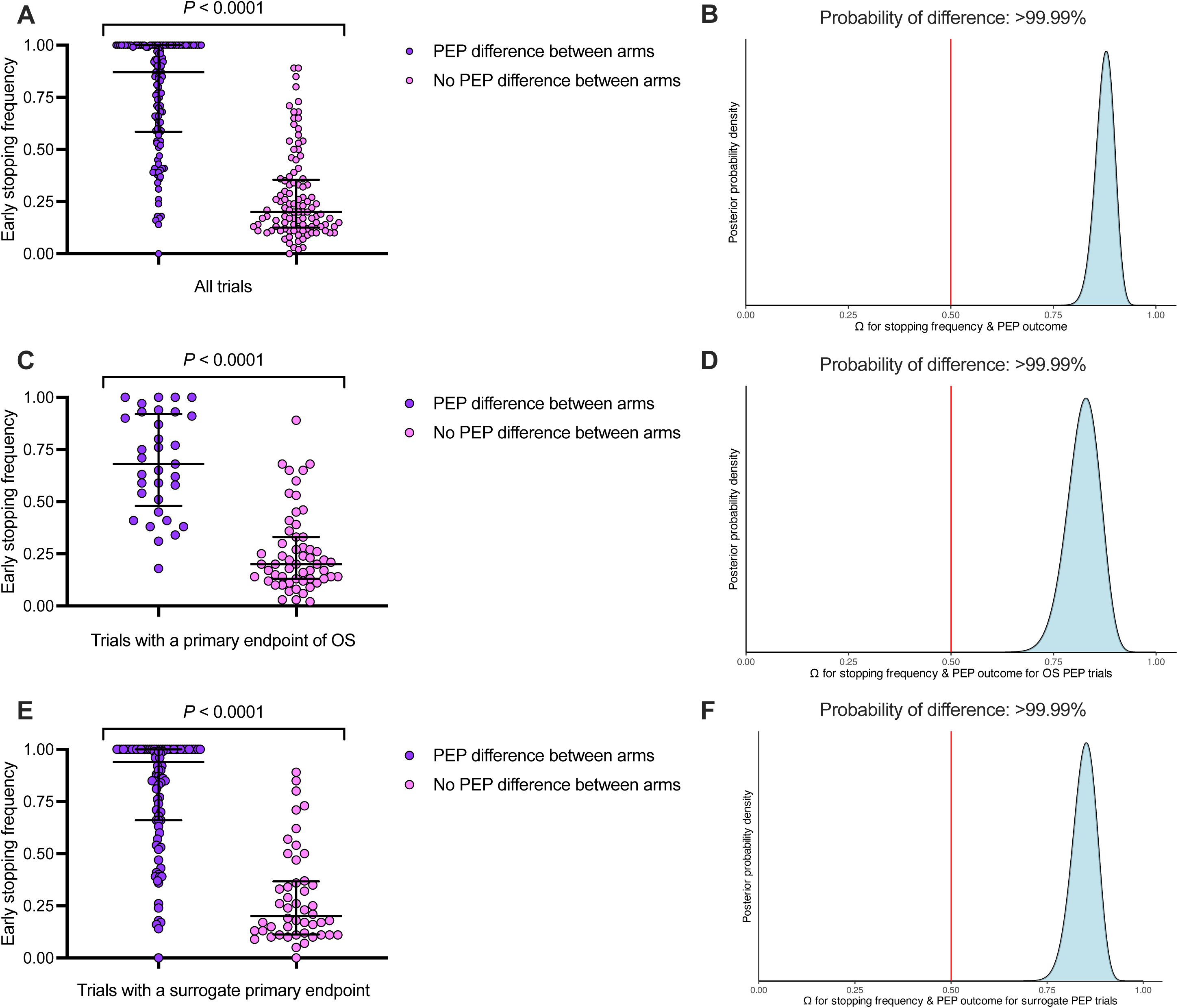
Comparison of early stopping frequency and the final published primary endpoint (PEP) outcome, defined as an efficacy difference between arms (efficacy or futility) or no difference. (A) Overall dataset and (B) corresponding posterior distribution; (C) trials with a PEP of overall survival (OS) and (D) corresponding posterior distribution; and (E) trials with a surrogate PEP and (F) corresponding posterior distribution. *P* by Mann-Whitney U, and posterior distribution by Bayesian Mann-Whitney U. Bars represent median and interquartile range. The red line represents the null effect Ω = 0.50.

Early stopping frequencies ≥ 0.75 had 96% sensitivity (95% CI, 91% to 98%) and 62% specificity (95% CI, 54% to 70%) for published PEP differences (**Figure S4**). With this criterion for recommending closure, the agreement between the Bayesian interim analysis and original analysis was high (Bayesian Cohen’s κ, 0.95; 95% CrI, 0.92 to 0.99) (**Figure 3A**). Overall, 82 trials (36%) had early stopping frequencies ≥ 0.75, including 16 trials evaluating OS (16 of 90, 18%) and 66 trials evaluating surrogate PEPs (66 of 140, 47%). Of these, 78 trials (95%) had demonstrated between-arm differences in the original PEP analysis. Further, of the 82 trials recommended for an earlier closure by Bayesian interim analysis, the majority (76%, 62 of 82) performed a frequentist interim analysis, and of these, 33 (40%) had already been closed early by the frequentist interim analysis. Regarding the remaining trials, 47 had published PEP differences and 101 trials did not.

**Figure 3.**
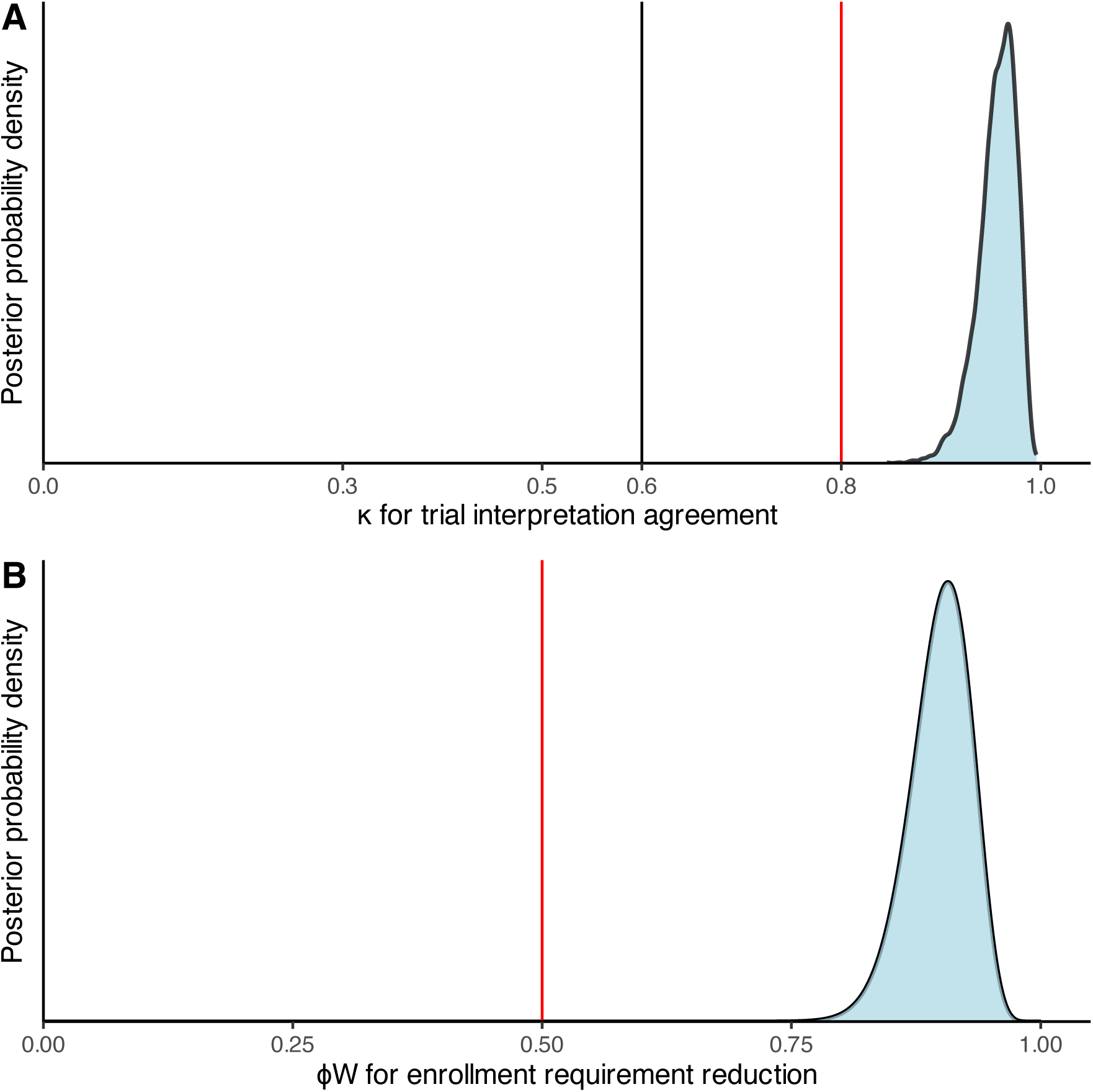
Bayesian interim analysis reduces enrollment requirements while preservation trial interpretation. (A) Posterior distribution of Bayesian Cohen’s κ, which indicates level of agreement between Bayesian interim analysis vs original analysis. κ < 0.6 indicates disagreement (black line), and κ > 0.8 indicates near-perfect agreement. (B) Posterior distribution comparing enrollment requirements between Bayesian interim analysis vs original analysis using Bayesian Wilcoxon signed-rank test. □_w_ = 0.50 (red line) indicates no difference between groups.

Two trial-level factors were associated with an early closure recommendation by the Bayesian interim analysis. Trials with an OS PEP were less likely to be recommended for early closure (aOR, 0.45; 95% CrI, 0.29 to 0.72; posterior probability of association, >99.99%) (**Figure S5**). On the other hand, industry funded trials were more likely to receive an early closure recommendation (aOR, 2.01; 95% CrI, 1.15 to 3.49; posterior probability of association, 99%) (**Figure S6**).

Using the median number of patients recommended for enrollment, Bayesian early stopping rules reduced the cumulative number of required participants by 20,543 (11%), or 89 patients averaged equally over all studied trials. The posterior probability that Bayesian interim analysis reduced enrollment requirements was >99.99% (median 490 patients vs 594 patients; _W_, 0.90; 95% CrI, 0.84 to 0.95; BF_10_, approaching ∞, by Bayesian Wilcoxon signed-rank test; *P* < 0.0001 by Wilcoxon signed-rank test) (**Figure 3B, S7**). The cumulative estimated cost savings from Bayesian early stopping rules was 851 million USD, or 3.7 million USD averaged equally per all studied trials.

## DISCUSSION

In the present large-scale study leveraging individual patient outcomes from 230 phase III trials, *in silico* Bayesian interim analysis reduced trial enrollment requirements without compromising overall trial interpretation compared with the original interim analysis strategy. Consistent with the notion that bias introduced by early stopping is limited, Bayesian interim analysis appeared to be highly sensitive for the final trial signal and did not lead to false-positive premature study closures.^35^ Minimum detectable effect thresholds may be a robust means of reliably identifying trials whose signal is strong enough for early closure. To our knowledge, the present study represents the largest endeavor of its kind, as prior studies have been limited to individual trials as case examples.^36–38^ As illustrated by this study, Bayesian frameworks facilitate a facile evaluation of multiple effect sizes beyond the null and provide a straightforward capacity to use separate priors for different hypotheses to strengthen model conclusions. Taken together, this study’s findings support the use of Bayesian interim analyses to improve the efficiency of randomized trials, lead to faster translation of new therapies to the clinic, reduced the overall resources and burden on patients required by phase III trials.

This study compared a single look *in silico* Bayesian interim analysis to the original published strategy, which often constituted multiple frequentist interim assessments. Multiple interim assessments may increase the odds of early stopping. Consequently, the present findings may be biased in favor of the original frequentist interim analysis strategy, and the efficiency gains from Bayesian interim analysis may be even greater with multiple looks. Notably, however, the approach to multiple Bayesian interim analyses and multiplicity corrections remains controversial.^39^ Some have argued that no multiplicity adjustment is necessary for Bayesian analyses, even for an unlimited number of looks, because Bayesian analyses do not condition on the null hypothesis, unlike frequentist analyses.^7, 12^ However, this argument is weakened if the prior distribution for the interim analysis is sequentially predicated on the posterior from the previous interim analysis.^39–41^ On the matter of prior selection, the priors used for the present study were default conventional distributions and not tailored to each study. Oncology-specific priors, trial-specific priors, or priors that are updated with the continuous availability of new trial data would probably lead to even greater advantages for Bayesian approaches than demonstrated in the present study.

In this study, an *in silico* Bayesian interim analysis was compared with the original analysis strategy employed by each trial, rather than an *in silico* frequentist interim analysis. A single *in silico* frequentist analysis would likely bias findings in favor of the Bayesian analysis by inadequately modeling real-life practices, which usually include multiple looks. This comparison would also raise the complication of whether to incorporate *in silico* alpha spending and multiplicity corrections for the frequentist analysis. Therefore, rather than adding uncertainty by attempting to model these parameters uniformly for all trials, we compared the Bayesian strategy to the strategy that was actually employed by each trial, which was notably trial-specific and thus a considerably more meaningful comparator than an *in silico* frequentist test. To this end, it is important to reiterate that 33 trials, which were closed early by the original frequentist interim analysis, were recommended for an even earlier closure by the Bayesian approach.

Consistent with expectations, underlying trial characteristics were associated with the probability of early stopping. Specifically, trials with surrogate PEPs were more likely to be recommended for early closure compared to trials with an OS PEP. Significant treatment effect sizes are arguably easier to obtain with composite surrogate endpoints, and in fact, the majority of regulatory drug approvals in oncology are on the basis of positive surrogate endpoints rather than OS.^42–44^ In light of these two observations, the strength of correlation between surrogate endpoints and OS has come under scrutiny, as well as the clinical meaningfulness of progression-based measures.^45–48^ Thus, an important caveat to this study is that most trials recommended for early closure used surrogate PEPs. In addition to PEP type, trials funded by industry sources were also associated with early stopping, which may be related to a complex combination of factors including endpoint selection, trial design, trial support, over-sampling and absence of optimism bias, and the generally observed overall higher rate of positive PEPs among industry-funded trials.^49–52^ Thus, unsurprisingly and consistent with the literature, the extent of potential efficiency gains from Bayesian interim analysis may be specific to the details of the experimental approach.

A key study limitation is that several assumptions used for the accrual simulations may not be accurate for all phase III trials, as accrual rate is a complex function of participating institutions, the size of the patient population, patients’ access to trial participation, competing studies, and other factors. While the MCID was chosen for representativeness and to improve the clinical meaningfulness of effect size assessment, it may not be appropriate for all scenarios. Furthermore, the MCID is a measure of relative, but not absolute, benefit, although relative measures of benefit remain the most common treatment effects evaluated in phase III trials.^26^ There are inherent limitations to reconstructed patient events from Kaplan-Meier curves, although we attempted to control these by excluding trials with poor quality reconstructions.^21^ The cost savings analysis used the median per-patient cost estimate from a separate study as a conservative estimate, although actual cost savings may be lower or higher depending on the specifics of the trial.^1, 2, 53^ Lastly, Bayesian interim analyses were not adjusted for patient-level prognostic covariates because individual patient-level data were obtained from unadjusted Kaplan-Meier curves. However, conditioning survival models with prognostic covariates is recommended for randomized trials to increase efficiency, and using adjusted Bayesian models in future studies may increase the efficiency gains beyond what is demonstrated here.^54, 55^

Taken together, this study supports the regular use of Bayesian interim analyses in the conduct of randomized trials. Bayesian interim analyses appear to reduce enrollment requirements while preserving the overall trial interpretation. Improved efficiency may translate to reduced costs, timely research reporting, reduced patient exposures to ineffectual treatments, and accelerated approval for effectual therapies. Future trials should consider formally incorporating Bayesian interim analyses into their pre-specified statistical analysis plan at the time of trial design.

## Data availability

Deidentified reconstructed individual patient-level data are available at the online repository figshare and accessible: https://figshare.com/articles/dataset/Reconstructed_survival_data_from_Phase_3_oncology_trials/26103268. The accrual simulation code for this study is provided in the Supplement.

## Acknowledgments

We thank Ann Sutton, Editing Services, Research Medical Library of The University of Texas MD Anderson Cancer Center, for editing the manuscript.

## Contributions

ADS, PM, TL, ZRM, and EBL conceptualized and designed the study. ADS, AM, TL, GK, JAJ, RK, MEA, RP, AK, CL, and EBL curated the data. ADS performed the formal analysis. EBL acquired funding. PM, TM, ZRM, and EBL supervised the study and provided study resources. ADS prepared the visualizations. All authors contributed to the interpretation of the data. ADS wrote the first draft, and all authors reviewed and revised the manuscript. All authors had full access to all of the data in the study and accept responsibility for submitting it for publication.

