## Supplement for "Bayesian Interim Analysis and Efficiency of Phase III Randomized Trials"

**Figure S1. Directed acyclic graph used to select confounders for Bayesian adjusted logistic regressions**

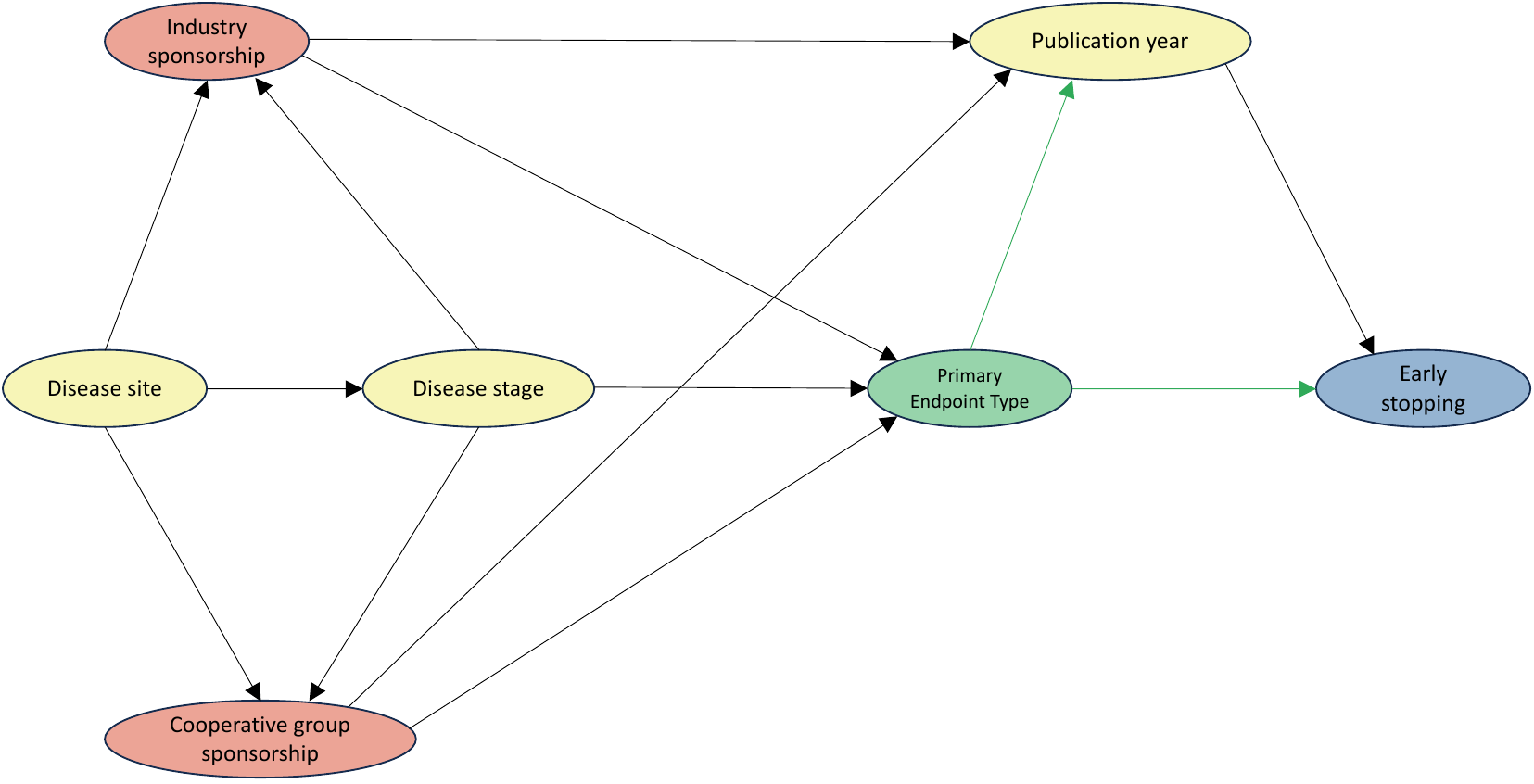

In this example, the outcome is shown in blue, the predictor in green, the confounders for the predictor in red, and the other variables in yellow.

**Figure S2. Trial selection flowchart**

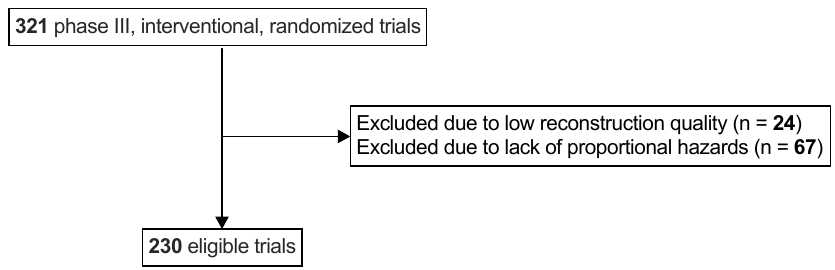

**Figure S3.** **Comparison of early stopping frequency and the primary endpoint (PEP) category, defined as overall survival (OS) or surrogate survival**

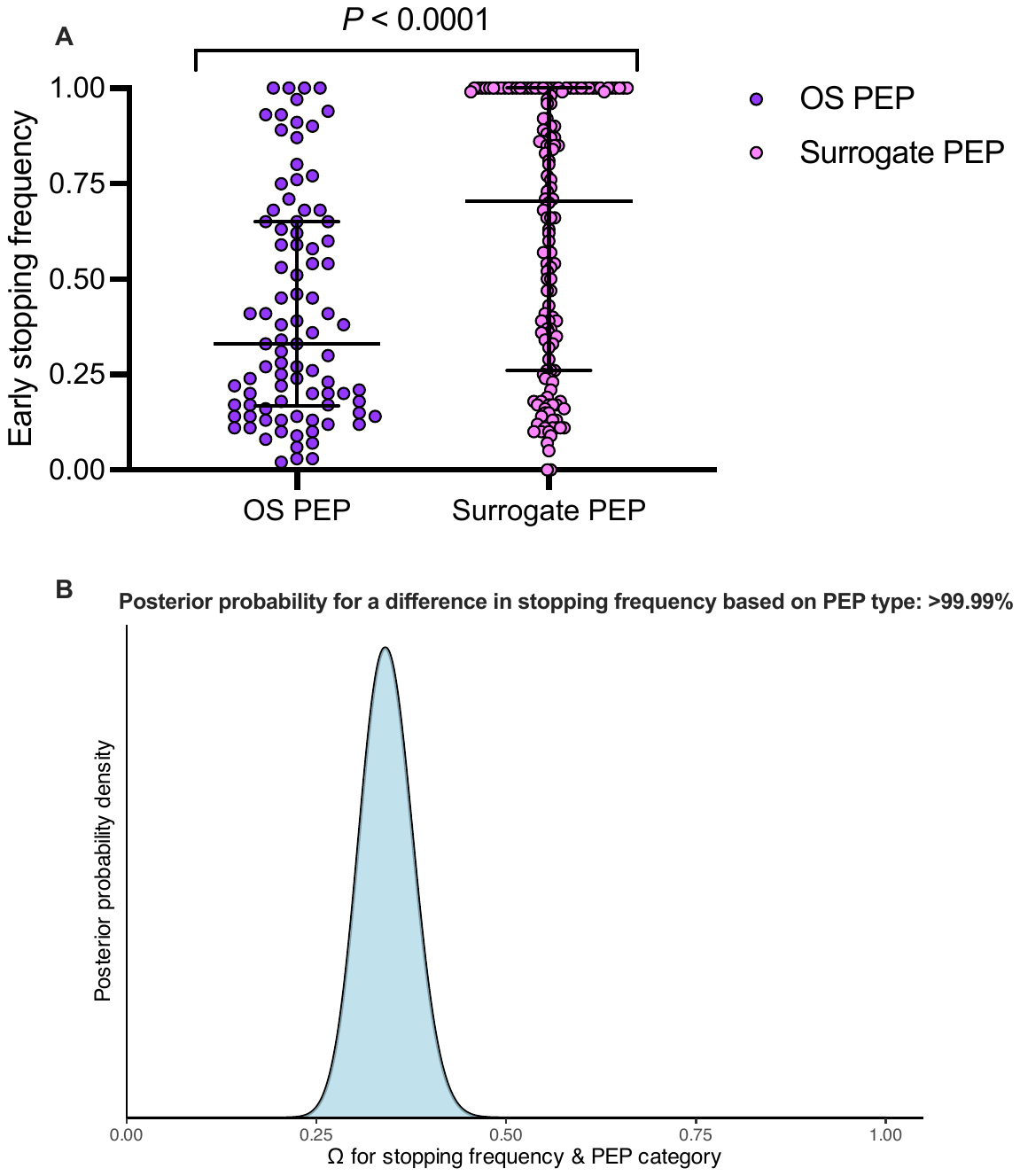

(A) Distribution per PEP type. *P* by Mann-Whitney U. Bars represent median and interquartile range. (B) Posterior distribution by Bayesian Mann-Whitney U. Ω = 0.50 represents the null.

**Figure S4. Receiver operating characteristic curve for early stopping frequency and published primary endpoint differences**

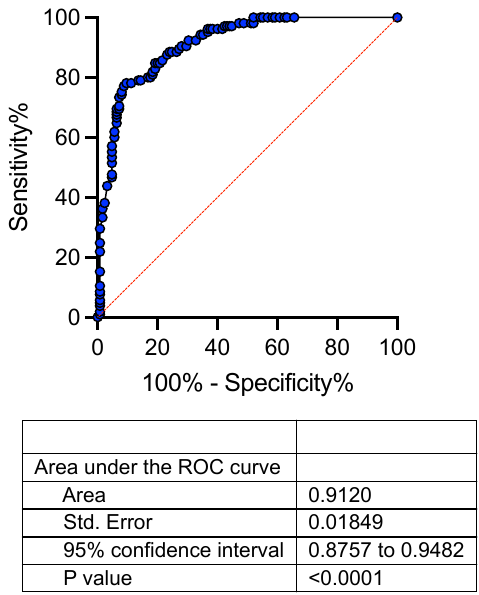

Abbreviations: ROC (receiver operating characteristic curve

**Figure S5. Posterior distribution for the association of primary endpoint (PEP) of overall survival (OS) vs surrogate and early stopping frequency ≥ 0.75**

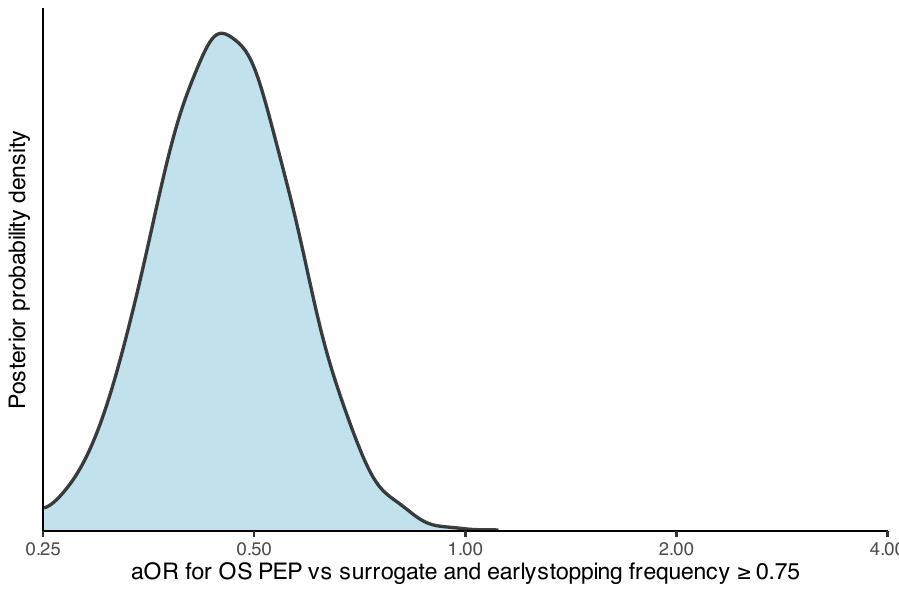

Bayesian logistic regression was adjusted for the confounders industry funding and cooperative group sponsorship. A skeptical prior was used. aOR = 1 reflects the null. Abbreviations: aOR (adjusted odds ratio).

**Figure S6. Posterior distribution for the association of industry sponsorship and early stopping frequency ≥ 0.75**

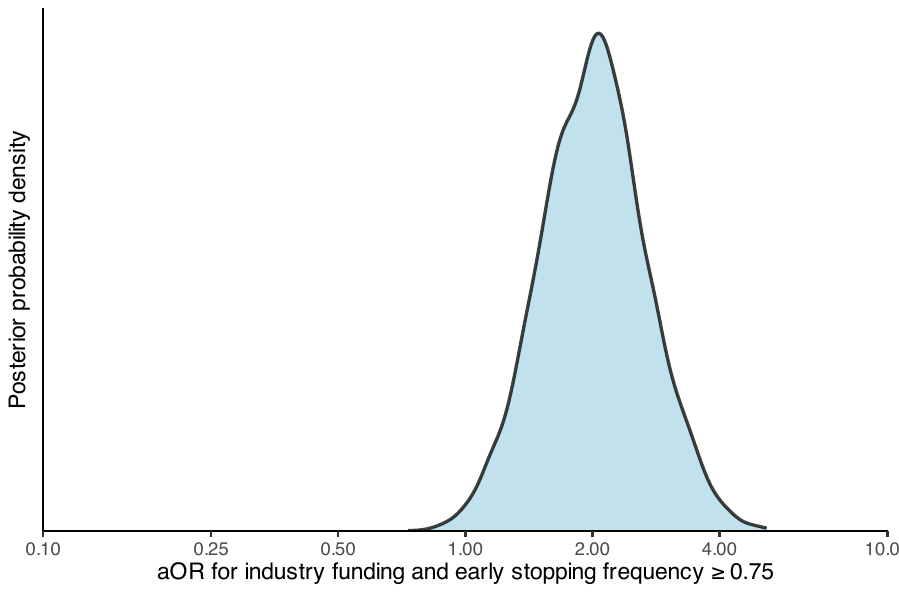

Bayesian logistic regression was adjusted for the confounders disease site and stage. A skeptical prior was used. aOR = 1 reflects the null. Abbreviations: aOR (adjusted odds ratio).

**Figure S7. Enrollment differences between the original analysis vs enrollment informed by Bayesian interim analysis**

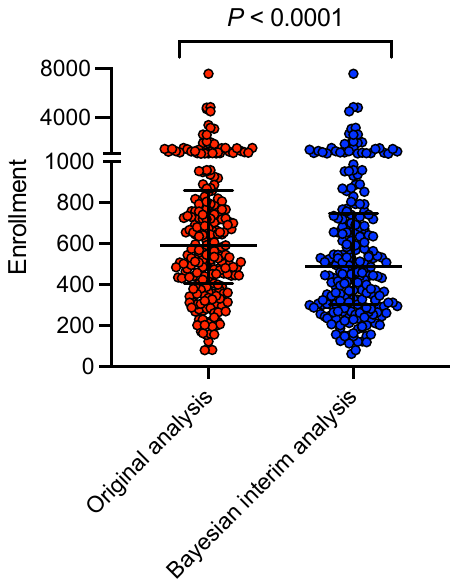

*P* value computed by Wilcoxon signed-rank test.

**Table S1**. **List of confounders for each predictor according to Figure S1**

| **Predictor** | **Confounder(s)** |
| --- | --- |
| Disease stage | Disease site |
| Disease site | None identified |
| Cooperative group sponsorship | Disease stage, disease site |
| Industry sponsorship | Disease stage, disease site |
| Year of publication | Primary endpoint type |
| Primary endpoint type | Industry sponsorship, cooperative group sponsorship |

### **Table S2. Characteristics of 230 included phase III oncology trials**

| **Characteristic*** | **Value** |
| --- | --- |
| Disease stage, n (%) |  |
| Solid M0 | 51 (22) |
| Solid M1 | 141 (61) |
| Hematologic | 38 (17) |
| Disease site |  |
| Breast | 45 (20) |
| Gastrointestinal | 41 (18) |
| Genitourinary | 35 (15) |
| Hematologic | 38 (17) |
| Thoracic | 40 (17) |
| Other** | 31 (13) |
| Treatment modality, n (%) |  |
| Systemic therapy | 224 (97) |
| Local therapy | 6 (3) |
| Cooperative group sponsorship, n (%) | 53 (23) |
| Industry sponsorship, n (%) | 202 (88) |
| Publication year, median (IQR) | 2015 (2013 to 2017) |
| Primary endpoint, n (%) |  |
| Overall survival | 90 (39) |
| Surrogate*** | 140 (61) |
| Frequentist interim analysis, n (%) | 170 (74) |
| Early closure | 64 (28) |
| Primary outcome, n (%) |  |
| Improved efficacy | 120 (52) |
| No improved efficacy | 105 (46) |
| Worse efficacy | 5 (2) |

Abbreviations: solid M0, non-metastatic solid tumor; solid M1, metastatic solid tumor; IQR, interquartile range.

*The primary endpoint of included trials was published between 2005 and 2020.

**Other disease sites included central nervous system, endocrine system, gynecologic system, head and neck, and skin.

***Examples of surrogate endpoints included progression-free survival, disease-free survival, and event-free survival.

### Script for accrual simulation

library(dplyr)

set.seed(123)

#df is the dataframe for the trial of interest with individual patient outcomes, including event_status_final and survival_time_final as well as treatment assignment

event_status_half <- sum(df$event_status_final) / 2

#set the accrual rate to be 5 to 15 patients enrolled per month

fastest_accrual <- 15

slowest_accrual <- 5

accrual_rate_monthly <- runif(1, min=slowest_accrual, max=fastest_accrual)

#Accrue the first month of patients

data_run <- df[sample(nrow(df), accrual_rate_monthly),]

data_run <- data_run %>% mutate(

surv_time_random = runif(n(), min = 0, max = 1),

surv_time_interim = ifelse(surv_time_final <= surv_time_random, surv_time_final, surv_time_random),

event_status_interim = ifelse(surv_time_final <= surv_time_random, event_status_final, 0)

)

events_interim <- sum(data_run$event_status_interim)

enrollment <- nrow(data_run)

enrollment_limit <- nrow(df)

enrollment_0.66 <- enrollment_limit * (2/3)

while (events_interim < event_status_half && enrollment < enrollment_0.66) {

variable_accrual <- runif(1, min=slowest_accrual, max=fastest_accrual)

#Randomly enroll more patients onto the trial if event_status goals are not met

add_patients <- df[sample(setdiff(seq_len(nrow(df)), rownames(data_run)), variable_accrual), ]

#Add survival times between 0 and the accrual time for this period for the new patients

add_patients <- add_patients %>% mutate(

surv_time_random = runif(n(), min = 0, max = 1),

surv_time_interim = ifelse(surv_time_final <= surv_time_random, surv_time_final, surv_time_random),

event_status_interim = ifelse(surv_time_final <= surv_time_interim, event_status_final, 0)

)

data_run <- data_run %>% mutate(

surv_time_interim = 1 + surv_time_interim,

)

data_run <- data_run %>% mutate(

surv_time_interim = ifelse(surv_time_final <= surv_time_interim, surv_time_final, surv_time_interim),

event_status_interim = ifelse(surv_time_final <= surv_time_interim, event_status_final, 0)

)

#Merge the existing patients with the newly accrued patients

data_run <- rbind(data_run, add_patients)

events_interim <- sum(data_run$event_status_interim)

enrollment <- nrow(data_run)

}
